# Perceived levels of mental health service accessibility and associated factors among psychiatric outpatients in Amanuel Mental Specialized Hospital, Addis Ababa, Ethiopia, 2024

**DOI:** 10.1101/2024.07.08.24310057

**Authors:** Surafel Ayenew, Kibrom Haile, Weldehawariat Leyew, Solomon Abate, Asrat Chaka

**Affiliations:** Department of Research and Training, Amanuel Mental Specialized Hospital, Addis Ababa, Ethiopia. Email: -; Department of Research and Training, Amanuel Mental Specialized Hospital, Addis Ababa, Ethiopia Email: -; Department of Psychiatry, Amanuel Mental Specialized Hospital, Addis Ababa, Ethiopia. Email: -

**Keywords:** Magnitude, Accessibility, barriers, Amanuel mental specialized hospital

## Abstract

**Background:** We live in multifaceted, dynamic and interconnected world in which the importance of physical, mental, and social wellbeing has not been well emphasized. Given the intricate nature of life, it is clear that mental health is essential to the welfare of people, communities, nations, and the entire planet. The main tactic to address unmet mental health needs has been to integrate psychiatric treatment into primary care, however this approach has been ineffective, and psychiatric services have been concentrated in Ethiopia and other major cities in sub-Saharan Africa. This study aimed to assess the perceived levels of mental health service accessibility and associated factors among psychiatric outpatients at Amanuel mental specialized hospital, Addis Ababa, Ethiopia, 2024.

**Objective:** To assess the perceived levels of mental health service accessibility and associated factors among psychiatric outpatients at Amanuel Mental Specialized Hospital, Addis Ababa, Ethiopia, 2024.

**Methods:** Institution-based cross-sectional study were employed from 03 January, to 03 February 2024. A total of 415 participants were participated. A systematic sampling technique was used. Binary logistic regression model was fitted to identify the factors associated with service accessibility. Adjusted odds ratio (AOR) with 95% confidence interval (CI) and p-value less than 0.05 during multivariable logistic regression were used to declare the factors associated with the outcome variable.

**Results:** The perceived level of mental health service accessibility was 10.4%; 95% CI: (7.5%, 25. 9%).Service accessibility was associated with urban resident [(AOR = 3.563, 95% CI: (1.065, 11.9155)], and Medication access [(AOR = 2.604, 95%CI: (1.220, 5.555)].

**Conclusions and recommendations:** The study observed that the prevalence of perceived service accessibility was found to be low in this study. Availability of medication and urban resident are the factors associated with perceived level of mental health service accessibility. There is a need to have proper coordination and cooperation among various mental health service providers.

## Introduction

Access to health care has a significant impact on overall health at all stages of life. With adequate use of health services, health problems can be detected and diagnosed earlier so that they can be treated more proactively. This, in turn has a positive effect on chronic diseases and life expectancy. Most healthcare systems around the world emphasize minimizing barriers to citizens’ access to healthcare, but many barriers systematically impede such a goal. Some important barriers are related to the fact that services are not sufficiently accessible, physically available, or reasonably priced(1). Lack of access to and use of mental health services remains particularly visible in low-and middle-income countries (2).

According to the World Health Organization (WHO), mental health is defined as a state of well-being in which each person realizes their potential, is able to cope with normal life stresses, is able to work productively and fruitfully, and is able to participate in their community (3). Thus, mental health is an integral part of health, including various activities that are directly and indirectly related to the promotion of mental well-being, the prevention and treatment of mental health disorders, and the rehabilitation of people with mental health problems (4). Although mental health disorders are known to be one of the greatest burdens of disability, with an estimated 37% of all healthy life years lost due to mental health disorders, it is recommended that mental health services be available at every possible level, mainly in communities and medical facilities(5).

Global burden of disease studies indicate that more than 5 percent of disability-adjusted life years (DALYs) and up to 15.7 percent of disability-adjusted lives (YLDs) worldwide are related to mental health problems (6,7). Despite the global upward trend in mental illness, this problem has not been properly treated as a physical illness and is largely neglected in many parts of the world, especially in low and middle-income countries (LMIC). This deficiency results in the limited, inadequate and inequitable provision of MH services(8), and would widen the care gap. This growing gap causes many health, social, and economic problems (9). Research has shown that the main barriers to providing MH services are information systems (10,11), financial and resource barriers(12), a lack of evidence-based policy and practice, and prevention, integration, and structural barriers(13).

In Ethiopia, neuropsychiatric disorders are estimated to account for 5.8 percent of the disease burden (14). Depression accounts for about 6.5 percent of illnesses (15). Mental health disorders accounted for about 11 percent of cases, mostly in rural areas. Schizophrenia and depression are among the ten most severe illnesses (16). It is often more difficult for people with mental health problems to access mental health services due to various attitudinal and structural barriers (7). To reduce treatment, many countries have taken the initiative to integrate mental health services into primary health care (17). Despite efforts to increase access to mental health services, treatment rates for mental illness remain low in LMICs. Low treatment rates may be due in part to a number of barriers to integration, including limited resources and low public demand for mental health services (18). Integrating psychiatric care with primary care, a key strategy to address unmet mental health needs, has remained inadequate, and psychiatric services are concentrated in large cities in sub-Saharan Africa. It is no different in Ethiopia, where the majority of care is provided by the Amanuel Mental Hospital, the country’s only psychiatric hospital, located in Addis Ababa. Despite the development of several regional psychiatric units, there is evidence of limited use of services in the wider community (19).Therefore, this study is aimed to assessing the levels of mental health services accessibility and associated factors among psychiatric outpatients of Amanuel mental specialized hospital.

## Method and materials

### Study area, design, and period

An institution based cross-sectional study was conducted from 03January to 03February 2024 in Amanuel Mental Specialized Hospital, Addis Ababa, Ethiopia. It is found in the capital city, which is the only specialized hospital that gives service for several years on mental health and related cases. The hospital has 239 beds including 11 private wing beds and 23 emergency beds, and also it has 13 outpatient departments (OPDs). On average, 10,320 patients receive the service per month, and the majority of patients stay in the hospital with their family members. The hospital is also playing a pivotal role as a continuous professional development center for psychiatric professionals of different levels.

### Population

All patients attending the psychiatric outpatient department of Amanuel mental specialized hospital were the source population. And all patients who attended at a psychiatric outpatient department, during the study period were the study population. All patients attending psychiatric outpatient with an age of 18 years and above were included in the study on the other hand, Patients who were critically ill and unable to respond were excluded.

### Sample size determination and sampling technique

The sample size was calculated using a single population proportion formula, taking into account the following assumptions:

Level of significance (α) =5% (at a confidence level of 95%), marginal error d=5% and P=0.5 (since there was no published research conducted in levels of mental health service accessibility). The Z value is 1.96 (n=sample size, P=proportion and d=marginal error). The sample size was 384. By considering 10% non-responses, the total sample size was 423.Systematic sampling technique was used to select representative samples of patients who had follow up at the outpatient department of Amanuel Mental Specialized Hospital. The first participant was selected by lottery method, then continued by every 23 intervals (k). The data were collected from every 23 other patients on follow up.

### Data collection techniques, variable measurement, and quality control

Structured Interviewer-administered questionnaire was used to collect the data. The questionnaire was prepared in English, translated to local language Amharic and translated back to English to ensure its consistency. The questionnaire includes socio-demographic characteristics, structural barriers, community barriers, provider/service user barriers, and healthcare related factors. Service accessibility was the dependent variable in this study and was operationally defined as services that are provided in a timely, continuous manner, develop close to the community and taking into account individual needs and preferences. Patients’ diagnosis and medication was taken from the patients’ medical record. We were also calculating Cronbach alpha (α) to check the reliability of measuring tool (VIF=0.97). One-day training on the basic techniques of data collection was given to data collectors and supervisors before the actual data collection period. Two Master of Science who had prior data collection experiences and knowledge of Amharic and Afan-Oromo language was recruited for data collectors and supervisors, respectively. Data collectors were also briefed on each item included in the data collection tool. The data collectors were observed daily, and the completed questionnaires were checked daily by the supervisor and the principal investigator. We pre-tested the questionnaire on 5% of the sample at Eka kotebe general hospital before the actual data collection period. The necessary modification was done based on the pre-test findings.

### Data processing and analysis

Coded and verified data were entered into a computer using EPI Data version 3.1 and imported into Statistical Package for Social Sciences (SPSS) Windows software version 21. Descriptive statistics such as frequency, percentage, mean, and median were calculated and presented using tables and graphics. Binary logistic analysis was performed to determine each explanatory variable, and variables with a p-value less than 0.25 in the bivariate analysis were entered into the multivariate analysis. A multivariate logistic regression analysis was performed to determine if there was statistically significant relationship between explanatory variables and outcome variables. P values less than 0.05 were considered statistically significant, and the strength of associations was reported as adjusted odds ratio with 95% C.I.

## Results

### Socio-demographic factors of respondents

A total of 415 respondents participated in this study yielding a response rate of 98%. Male study participants accounted for 58.6 %. The majority (38.6%) of the respondents were 26-35 years old. Most (47%) of study participants were married by marital status. Regarding the residents, 88.2% of the respondents were from urban. Of the total study participant, 29.6% participants were private employees (Table 1).

**Table 1.** Socio-demographic characteristics of psychiatric outpatients in Amanuel mental specialized hospital, Addis Ababa, Ethiopia 2024 (n= 415)

| Variables | Category | Frequency | Percentage |
| --- | --- | --- | --- |
| Sex | Male | 243 | 58.6% |
|  | Female | 172 | 41.4% |
| Age category | <26 | 108 | 26% |
|  | 26-35 | 160 | 38.6% |
|  | >35 | 147 | 35.4% |
| Marital status | Married | 195 | 47% |
|  | Single | 147 | 35.4% |
|  | Divorced | 30 | 7.2% |
|  | Widowed | 35 | 8.4% |
|  | Separated | 8 | 1.9% |
| Occupation | Unemployed | 88 | 21.2% |
|  | Private employee | 123 | 29.6% |
|  | Farmer | 34 | 8.2% |
|  | Government employee | 93 | 22.4% |
|  | Daily Laborer | 27 | 6.5% |
|  | Others | 50 | 12% |
| Resident | Urban | 366 | 88.2% |
|  | Rural | 49 | 11.8% |

### Perceived level of mental health Services accessibility

The perceived level of mental health service accessibility was 43(10.4%; 95 %CI: 7.5, 13.5). Among the study participants, 89.6% had perceived level of service non-accessibility.

### Community factors

Among the respondents (71.3%) had received stigma from the community. Regarding the way; the community treated mentally ill patients, 288(69.4%) were maltreated by the community. The majority of the respondents, 216(52%), were lived in communities with low economic status (Table 2.)

**Table 2.** Community factors affecting perceived level of service accessibility among psychiatric outpatients in Amanuel mental specialized hospital, Addis Ababa, Ethiopia 2024 (n= 415)

| Variables | Category | Frequency | Percentage |
| --- | --- | --- | --- |
| Stigma | Yes | 296 | 71.3% |
|  | No | 119 | 28.7% |
| Mal-treatment | Yes | 288 | 69.4% |
|  | No | 127 | 30.6% |
| Support structure | Yes | 187 | 45.1% |
|  | No | 228 | 54.9% |
| Economic status | Low | 216 | 52% |
|  | Medium | 175 | 42.2% |
|  | High | 24 | 5.8% |
| Traditional healers | Yes | 110 | 26.5% |
|  | No | 305 | 73.5% |
| Board Support | Yes | 202 | 48.7% |
|  | No | 213 | 51.3% |

### Health facility factors

The majority 341(82.2%) of the study participants reported that the health facilities lack capacity to give mental health services. Among the participants, 357(86%) patients perceived that their nearby health facilities lack medications to treat mental illness (Table 3).

**Table 3.**
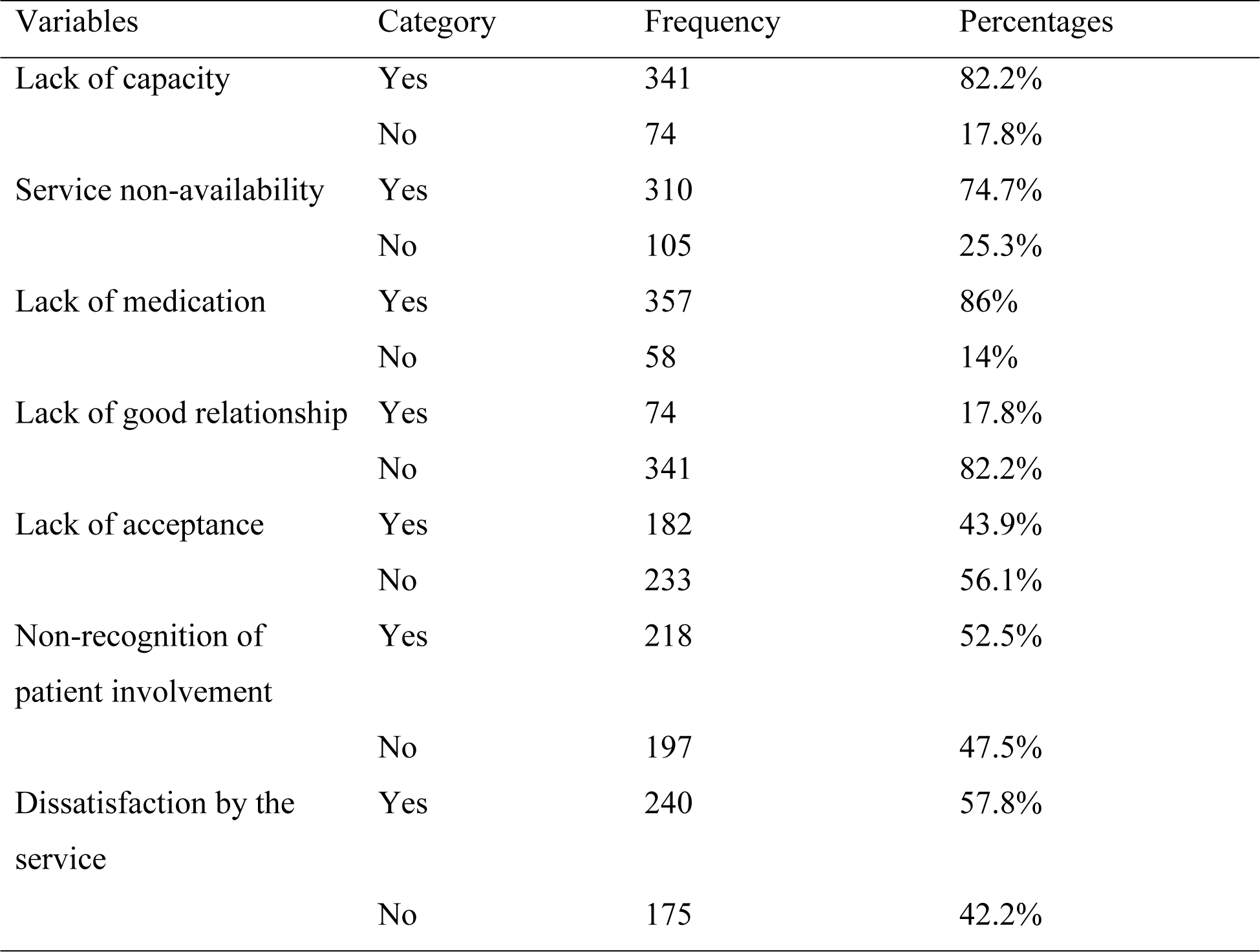
Health facility factors affecting perceived level of service accessibility among psychiatric outpatients in Amanuel mental specialized hospital, Addis Ababa, Ethiopia 2024 (n= 415)

### Structural factors

The result of this study showed that most (56.6%) of the participants perceived that there was structural stigma for mental health. About 73.5% of the respondents were lack services accessibility due to lack of priority for mental health (Table 4).

**Table 4.** Structural factors affecting perceived level of service accessibility among psychiatric outpatients in Amanuel mental specialized hospital, Addis Ababa, Ethiopia 2024 (n= 415)

| Variables | Category | Frequency | Percentage |
| --- | --- | --- | --- |
| Structural stigma | Yes | 235 | 56.6% |
|  | No | 180 | 43.4% |
| Lack of priority | Yes | 305 | 73.5% |
|  | No | 110 | 26.5% |
| Low scale-up | Yes | 345 | 83.1% |
|  | No | 70 | 16.9% |

### Provider/user factors

Of the respondents 285(71.1%) of them reported that they did not visit there nearby health facility due to lack of patient involvement in the treatment process. Majority 53.7% of the respondents did not access mental health services because of self-stigma. (Table 5)

**Table 5.** Provider/user factors affecting perceived level of service accessibility among psychiatric outpatients in Amanuel mental specialized hospital, Addis Ababa, Ethiopia 2024 (n= 415)

| Variables | Category | Frequency | Percentage |
| --- | --- | --- | --- |
| Lack of involvement | Yes | 295 | 71.1 |
|  | No | 120 | 28.9 |
| Self- stigma | Yes | 223 | 53.7 |
|  | No | 192 | 46.3 |
| Lack of self esteem | Yes | 227 | 54.7 |
|  | No | 188 | 45.3 |
| Lack of self-help group | Yes | 263 | 63.4 |
|  | No | 152 | 36.6 |

### Factors associated with perceived mental health service accessibility

We did binary logistic regression analysis in order to find the candidate variables for multivariable logistic regression analysis. And variables with p-value less than or equal to 0.25 were fitted to multivariable logistic regression analysis. In the bi-variable logistic regression analysis; sex, Resident, Stigma, Maltreatment, Traditional healers, lack of capacity, service non-availability, lack of medication, lack of good relationship, lack of acceptance, lack of patient involvement, dissatisfaction, low scale-up and self-stigma were found to be a candidate variable for multivariable logistic regression. In the final model, factors significantly associated with perceived service accessibility were resident and medication access. While variables like age, marital status, occupation, support structure, economic status of the community, board support, structural stigma, lack of priority, national policy, lack of patient involvement, lack of self-esteem, lack of self-help group, were not significantly associated with service accessibility.

In multivariable logistic regression analysis, Resident and Medication were significantly associated with perceived service accessibility. The odds of perceived mental health service accessibility among service users of urban residents were **3.563 (AOR: 3.563, 95**%CI: **1.065, 11.915)** higher compared with Rural residents.

The odds of perceived service accessibility among respondents who had medication access were **2.604** times **[(AOR=2.604, 95% CI: (1.220, 5.555)]** higher compared with counterparts(Table 6)

**Table 6.** Factors associated with perceived level of mental health services accessibility among psychiatric outpatients in Amanuel Mental Specialized Hospital 2024(n=415)

| Variables | Category | Service-accessibility |  | COR(95% CI) | AOR(95% CI) |
| --- | --- | --- | --- | --- | --- |
|  |  | Yes | No |  |  |
| Sex | Male | 21 | 222 | 1 | 1 |
|  | Female | 22 | 150 | 1.550(0.823,2.919) | 1.130(0.507,2.253) |
| Resident | Urban | 40 | 296 | 3.423(1.031,11.366) | **3.563(1.065,11.915)\*** |
|  | Rural | 3 | 76 | 1 | 1 |
| Stigma | Yes | 27 | 268 | 0.655(0.339,1.265) | 1.152(0.508,2.614) |
|  | No | 16 | 104 | 1 | 1 |
| Mal treatment | Yes | 27 | 256 | 0.765(0.397,1.474) | 0.889(0.268,2.948) |
|  | No | 16 | 116 | 1 | 1 |
| Traditional Healers | Yes | 8 | 104 | 1.698(0.762,3.781) | 1.326(0.572,3.077) |
|  | No | 35 | 268 | 1 | 1 |
| Lack of capacity | Yes | 29 | 306 | 0.447(0.224,0.892) | 0.976(0.156,6.126) |
|  | No | 14 | 66 | 1 | 1 |
| Service non-availability | Yes | 26 | 284 | 0.474(0.246,0.914) | 1.080(0.333,3.504) |
|  | No | 17 | 88 | 1 | 1 |
| Availability of medication | Yes | 31 | 326 | 0.365(0.175,0.760) | **2.604(1.220,5.555)\*** |
|  | No | 12 | 46 | 1 | 1 |
| Lack of good relationship | Yes | 4 | 70 | 0.442(0.153,1.279) | 1.692(0.570,5.026) |
|  | No | 39 | 302 | 1 | 1 |
| Lack of acceptance | Yes | 15 | 173 | 0.616(0.319,1.192) | 1.024(0.421,2.491) |
|  | No | 28 | 199 | 1 | 1 |
| Patient involvement | Yes | 18 | 200 | 0.619(0.327,1.173) | 0.945(0.410,2.177) |
|  | No | 25 | 172 | 1 | 1 |
| Dissatisfaction | Yes | 20 | 220 | 0.601(0.319,1.32) | 1.062(0.467,2.415) |
|  | No | 23 | 152 | 1 | 1 |
| Low scale-up | Yes | 33 | 312 | 0.635(0.297,1.356) | 1.517(0.696,3.307) |
|  | No | 10 | 60 | 1 | 1 |
| Self-stigma | Yes | 18 | 205 | 0.587(0.309,1.112) | 0.845(0.365,1.954) |
|  | No | 25 | 167 | 1 | 1 |

Note: - \*- Significantly associated at p<0.05
1- Reference
AOR = Adjusted odds ratio; COR = crude odds ratio; CI = confidence interval

## Discussion

The present study was conducted in Amanuel Mental Specialized Hospital, and it presents mental health service users’ perception of service accessibility and the barriers to utilizing mental health services. Accordingly, the perceived level of mental health service accessibility among mental health service users in Amanuel Mental Specialized Hospital was 10.4%. The predictors of perceived service accessibility in this study included Resident and Medication availability.

The perceived level of mental health service accessibility among patients attending outpatient departments in Amanuel Mental specialized hospital was 10.4%. This data was much lower than the national healthcare access and quality index for Ethiopia 2019 (33 %) that was presented by the national data management center for health (NDMC). This could be due to mental health care service barriers including lack of mental health policy, limited professionals and financial resources, stigma and discrimination towards mental health problems and its treatment.

This finding revealed that the odds of perceived service accessibility among urban residents were almost four times higher compared to rural residents. The findings suggest that a large proportion of patients travel long distances to access mental health care, primarily located in the country’s capital, Addis Ababa, and this may prevent many people from accessing mental health services. This finding is consistent with previous research in Nigeria, which showed that long waiting times and long distances to access mental health services are barriers to service use, especially among rural populations (20). Another study showed that depressed patients living 30 to 60 miles away from psychiatric health services were less likely to receive therapy compared to those who were living within shorter distances(21). Another study found that depressed patients who lived 30 to 60 miles from psychiatric health services were less likely to receive treatment than those who lived a shorter distance(22)

In the present study, availability of medication was significantly associated with perceived mental health service accessibility; the odds of perceived mental health service accessibility among respondents who had medication access were nearly three times more likely compared to counterparts. Offering free psychiatric medication to psychiatric patients and their inclusion in the health insurance was recommended by some psychiatrists as a way of increasing mental health services utilization and access in Sudan. Health insurance coverage as a successful policy to improve patient’s access to psychiatric medications was suggested by policymakers in the previous study in Sudan(23). Furthermore, increasing the availability of essential psychiatric medications was recommended by the WHO(24) to strength mental health system in Sudan. This is not different in Ethiopia. The Ethiopian government implements community based health insurance system to increase the utilization of health services and reduced the incidence of catastrophic health expenditure.

## Strength and Limitations of the Study

The study addressing adequate sample size. To the best of the researcher’s knowledge this is the first study to assess the perceived service accessibility among mental health service users were the strength of the study. The study has been restricted only to mental health service users in Amanuel Mental Specialized Hospital. Since this is a cross-sectional study it does not show the casual relationship between service accessibility and mental health was the limitations of this study.

## Conclusions

The study observed that the prevalence of perceived mental health service accessibility was found to be low in this study. Availability of medication and urban resident are the factors associated with perceived level of mental health service accessibility. There were many barriers to utilizing the services. There is still lack of awareness of mental health and mental health issues, and services have not yet reached those in need. The service needs to reach the communities, especially the remote areas. Appropriate coordination and collaboration between different mental health service providers is necessary. To improve the utilization of mental health services, a periodical evaluation must be carried out.

## Data Availability

All the data supporting the results are within the manuscript. Additional detailed information and raw data are available from the corresponding author on logical request

## Acronyms and Abbreviations

AOR: Adjusted Odds Ratio
CI: Confidence Interval
COR: Crude Odds Ratio
DC: Data Collector
DALYs: Disability Adjusted Life Years
ETB: Ethiopia Birr
LMICs: Low and Middle Income Countries
MH: Mental Health
OPD: Out Patient Department
PI: Principal Investigator
SPSS: Statistical Package for the social sciences
WHO: World Health Organization
YLDs: Years Lived with Disability

## Declarations

### Ethics approval and consent to participations

Ethical approval was obtained from Ethical Review Board of Amanuel Mental Specialized Hospital. Written informe
d consent was obtained from each participant. The natures of the study was explained, which required answering of questions and no invasive procedure or associated risks were involved. They were also having informed of opting out of the study at any point without any penalty, should they decide to leave. Anonymity and confidentiality of information obtained was assured and maintained.

### Consent for publication

Not applicable

### Availability of data and materials

“All the data supporting the results are within the manuscript. Additional detailed information and raw data are available from the corresponding author on logical request. ”

### Authors’ contributions

“S.A. and K.H. conduct study design, data collection, data analysis and manuscript preparation and W.L. and S.A.perform study design and manuscript preparation and A.C.wrote the main manuscript text. Finally, all authors reviewed, finalized, and approved the manuscript.”

## Acknowledgements

The authors would like to thank Amanuel Mental Specialized Hospital for its technical support. We are also extended our appreciation to the study participants and data collectors.

## Authors’ information

1. SA, Department of research and training, Amanuel Mental Specialized Hospital, Addis Ababa, Ethiopia
2. KH, Department of research and training, Amanuel Mental Specialized Hospital, Addis Ababa, Ethiopia
3. WL, Department of research and training, Amanuel Mental Specialized Hospital, Addis Ababa, Ethiopia
4. SA, Department of research and training, Amanuel Mental Specialized Hospital, Addis Ababa, Ethiopia
5. AC, Department of psychiatry, Amanuel Mental Specialized Hospital, Addis Ababa, Ethiopia

